# A blueprint for biobanking in everyday clinical practice in psychiatry: The Munich Mental Health Biobank

**DOI:** 10.1101/2022.05.05.22274583

**Authors:** JL Kalman, G Burkhardt, K Adorjan, BB Barton, S De Jonge, D Eser-Valeri, CM Falter-Wagner, U Heilbronner, A Jobst, D Keeser, C Koenig, G Koller, N Koutsouleris, C Kurz, D Landgraf, K Merz, R Musil, AM Nelson, F Padberg, S Papiol, O Pogarell, R Perneczky, F Raabe, MA Reinhard, A Richter, T Rüther, MS Simon, A Schmitt, L Slapakova, N Scheel, C Schüle, E Wagner, SP Wichert, P Zill, P Falkai, TG Schulze, EC Schulte

## Abstract

Translational research on complex, multifactorial mental health disorders, such as bipolar disorder, major depressive disorder, schizophrenia, and substance use disorders requires databases with large-scale, harmonized, and integrated real-world and research data.

The Munich Mental Health Biobank (MMHB) is a mental health-specific biobank that was established in 2019 to collect, store, connect, and supply such high-quality phenotypic data and biospecimens from patients and study participants, including healthy controls, recruited at the Department of Psychiatry and Psychotherapy and the Institute of Psychiatric Phenomics and Genomics, University Hospital of the Ludwig-Maximilians-University (LMU), Munich, Germany. Participants are asked to complete a questionnaire that assesses sociodemographic and cross-diagnostic clinical information, provide blood samples, and grant access to their existing medical records. The generated data and biosamples are available to both academic and industry researchers. In the current manuscript, we outline the workflow and infrastructure of the MMHB, describe the clinical characteristics and representativeness of the sample collected so far, and reveal future plans for expansion and application.

As of October 31, 2021, the MMHB contains a continuously growing set of data from 578 patients and 104 healthy controls (46.37% female; median age, 38.31 years). The five most common mental health diagnoses in the MMHB are recurrent depressive disorder (38.78%; ICD-10: F33), alcohol-related disorders (19.88%; ICD-10: F10), schizophrenia (19.69%; ICD-10: F20), depressive episode (15.94%; ICD-10: F32), and personality disorders (13.78%; ICD-10: F60). Compared with the average patient treated at the recruiting hospitals, MMHB participants have significantly more mental health-related contacts, less severe symptoms, and a higher level of functioning. The distribution of diagnoses is also markedly different in MMHB participants compared with individuals who did not participate in the biobank.

After establishing the necessary infrastructure and initiating recruitment, the major tasks for the next phase of the MMHB project are to improve the pace of participant enrollment, diversify the sociodemographic and diagnostic characteristics of the sample, and improve the utilization of real-world data generated in routine clinical practice.

## 1. Background

In recent years, technological and methodological advances have enabled diseases to be studied at an unprecedented depth and expanse. Biobanks, which collect, store, manage, and share large sets of biosamples and deep phenotyping information, are critical infrastructures for the clinical translation of these advances into patient care.

Establishing such rich datasets is especially important for mental health disorders because current diagnostic categories have low prognostic value, disease biomarkers are largely lacking, and novel therapeutic approaches have often failed to reduce the global burden of these disorders (1,2). Nevertheless, mental health disorders are underrepresented in population-based biobanking efforts, and even the largest and most successful biobanks, such as the UK Biobank and the Estonian Biobank, began to collect extensive mental health-related phenotypes only years after their initiation (3,4). The reasons for this delay may include the significant resources and commitment required for mental health phenotyping and the difficulties in utilizing real-world data for mental health research because of the lack of data structure, standardization, and interoperability (5). Social stigma attached to many mental disorders and the special legal implications, such as legal guardianship, may further reduce the willingness and ability of patients to participate in biobanking efforts.

Similar trends are seen in Germany: Currently, only a few, large mental health facilities routinely collect phenotype and biological data, and most of these efforts focus on individuals participating in clinical studies and do not include patients in routine clinical practice (6–8). Therefore, to facilitate biobanking in mental health, we established the Munich Mental Health Biobank (MMHB), which intimately entwines the collection of both routine clinical and research phenotyping data and biological specimens at a large tertiary care center in Munich, Germany, comprising the Department of Psychiatry and Psychotherapy (DPP) and the Institute of Psychiatric Phenomics and Genomics (IPPG). In the current manuscript, we describe the workflow and information technology infrastructure of the MMHB, provide a first glimpse into the clinical characteristics of the sample collected so far, analyze the representativeness of the sample with regard to a variety of key clinical and sociodemographic characteristics, and reveal future plans for expansion and application.

## 2. The MMHB

The DPP and IPPG are part of the hospital system of the University Hospital of the Ludwig-Maximilians-University (LMU) Munich, Germany. With 3222 unique cases (inpatient: 1058; day hospital: 130; and outpatient: 2034) annually (2020 data), these two institutes are among the largest university-based mental health facilities in Germany. Both institutes are internationally renowned strongholds of mental health research and are currently hosting over 43 ongoing clinical studies. However, until recently, neither the DPP nor the IPPG had access to an organized, standardized, high-throughput hospital-wide collection of phenotypes and biosamples, markedly limiting large-scale translational research. This changed in 2019 with the establishment of the MMHB. Since then, all individuals treated at the DPP and IPPG, as well as study participants (both patients with a mental health diagnosis and healthy controls with no self-reported lifetime mental health diagnosis participating in clinical or non-clinical trials), are eligible for inclusion in the MMHB and can be invited to provide informed consent to the collection of phenotypic information and biosamples as part of the biobanking effort.

### 2.1. Informed Consent

For the following activities, the MMHB aims to obtain modular informed consent from participants that conforms with the General Data Protection Regulation of the European Union: 1) the collection, storage, analysis, scientific utilization, and distribution of deep phenotypic data and biosamples in a double-pseudonymized form to academic and non-academic research institutions, national and international data archives, and companies, and the linkage of this data to routine clinical data; 2) re-contacting participants to request additional data and biosamples and/or to invite them to participate in new studies; 3) re-contacting participants to seek consent for linking the collected data to other databases; and 4) re-contacting participants to inform them about actionable incidental findings. By providing their consent, MMHB participants allow researchers to use their phenotypic data and biosamples for any type of research, including omics studies and the creation of induced pluripotent stem cell (iPSC) lines, for 30 years. MMHB participants can withdraw their consent at any time without providing a specific reason. In case of withdrawal of consent, they can decide whether their data and biosamples should be deleted and destroyed or anonymized.

### 2.2. The MMHB phenotyping battery

The MMHB phenotyping battery consists of three components: 1) a ***basic phenotyping module***, which builds on previous national multi-centric phenotyping efforts, such as the PsyCourse Study and the PD-CAN study; 2) a module consisting of seven ***standardized self-rating measurements*** with cross-disorder utility; and 3) a ***variable module*** that highlights a specific topic of research in more detail and changes every two years *(Table 1)* (9). In 2020 and 2021, the variable module focused on metabolic risk, eating behavior, and stress related to the COVID-19 pandemic. To reduce the biobanking-associated clinician workload, self-rather than observer-rating tools are used to obtain patient characteristics and symptom scores. The MMHB phenotyping battery is complemented by data collected in association with specific scientific projects or clinical trials.

**Table 1:** Overview of the phenotypes assessed with the Munich Mental Health Biobank (MMHB) phenotyping battery

| Module | Questionnaire | Reference |
| --- | --- | --- |
| Basic phenotyping | Sociodemographic information |  |
|  | Psychiatric medical history |  |
|  | Family history |  |
|  | Pharmacologic treatment |  |
|  | Self-destructive behavior & suicidality |  |
|  | Substance abuse, including the Alcohol Use Disorders Identification Test (AUDIT-C) | Bush et al. 1998 (10) |
| Standardized self-rating instruments | Childhood Trauma Screener | Grabe et al. 2021 (11) |
|  | Brief Resilience Scale | Smith et al. 2008 (12) |
|  | UCLA Loneliness Scale (3-item version) | Hughes et al. 2004 (13) |
|  | Lubben Social Network Scale | Lubben et al. 2006 (14) |
|  | WHO-5 Well-Being Index | Topp et al. 2015 (15) |
|  | Patient Health Questionnaire (PHQ-9) | Kroenke et al. 2001 (16) |
|  | Munich Chronotype Questionnaire | Roenneberg et al. 2003 (17) |
| Variable module (2020/2021) | Questions on metabolic risk | Adapted from Barton et al. 2020 (18) |
|  | Questions on eating behavior | Adapted from Cappelleri et al. 2009 (19) |
|  | COVID-19 Pandemic Mental Health Questionnaire (CoPaQ) | Rek et al. 2021 (20) |

### 2.3. Biosamples

A limited amount of blood (maximum 70 mL) and, if available, cerebrospinal fluid (maximum 10 mL) is collected from consented individuals during routine clinical examinations or study visits *(Supplementary Table S2)*. Participants may also be asked to provide saliva, hair, and stool samples.

### 2.4. Linked routine clinical data

Routine clinical data stored in electronic health records or other databases can be linked with the phenotypic information and biosamples collected from MMHB participants and then used for research purposes.

### 2.5. Data and sample processing

The collected phenotypic data and biosamples are processed by specially trained staff according to standard operating procedures for data collection, handling, and storage. For each MMHB participant, an electronic record, which includes general and disease-specific clinical information, dates of phenotype and biosample acquisitions, and information on sample processing (i.e., identification code, quantity, location of storage, and type of preparation) is captured in CentraXX (Kairos GmbH, Bochum, Germany), a standardized laboratory information management system. CentraXX is fully integrated into the laboratory automation infrastructure and, besides the administration of biosamples, enables the acquisition and management of all clinical and phenotypic data of participants. Biosamples (DNA, RNA, serum, plasma, etc.) are processed on site with automated systems (ChemagicStar, Decapper/Barcode Reader, and a StarPlus System; Hamilton Robotics GmbH, Martinsried, Germany) and are stored at −80°C in an automated sample storage and management system (BIOS M System, Hamilton Storage GmbH, Bonaduz, Switzerland) *(Figure 1)*.

**Figure 1:**
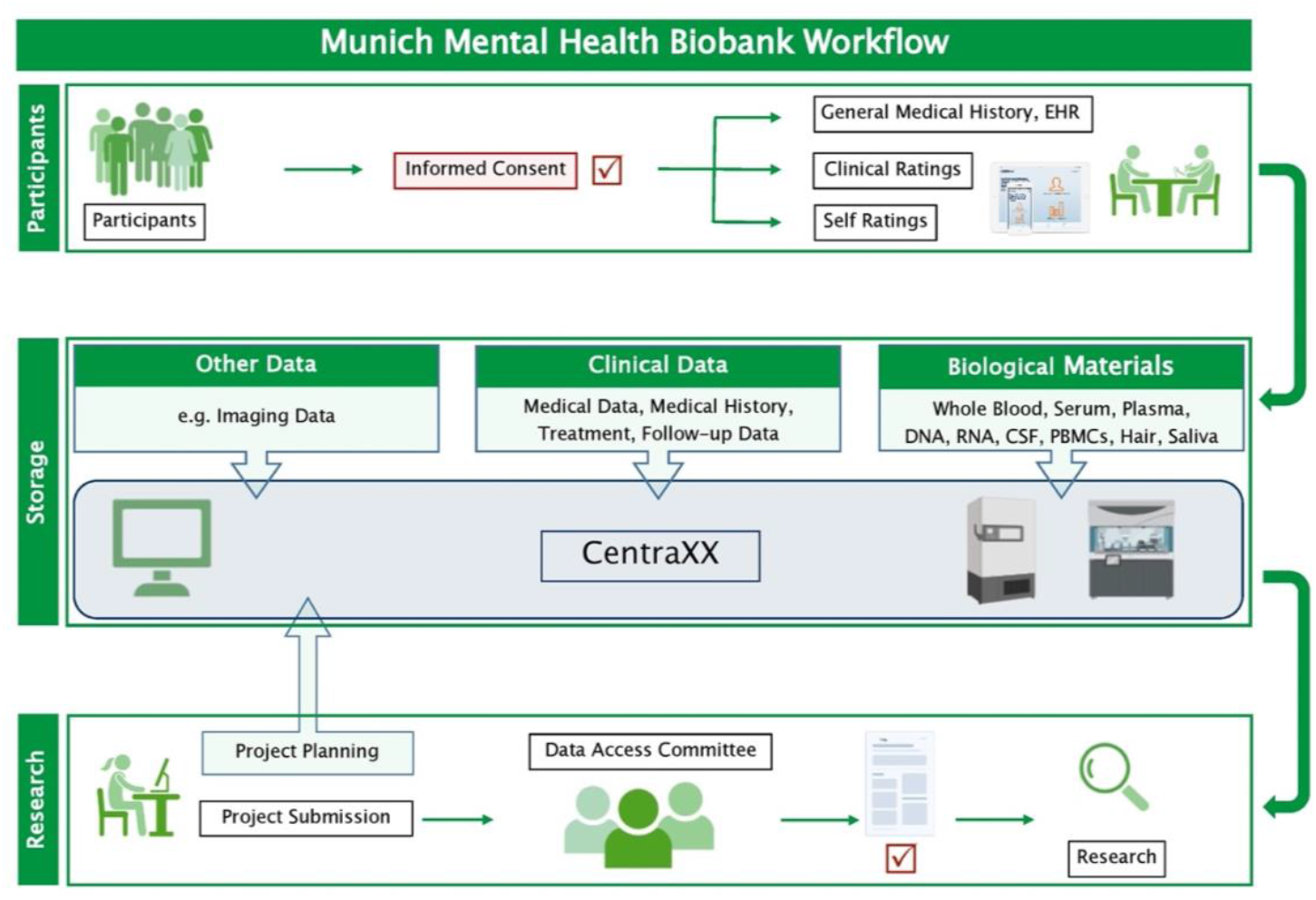
Workflow of the Munich Mental Health Biobank. Clinical data and biological materials are stored in pseudonymized form in the information management system CentraXX (Kairos GmbH, Bochum, Germany). Biological materials are stored at −80°C, except for peripheral blood mononuclear cells, which are stored in liquid nitrogen *(Supplementary Table S2)*. Researchers can request data access by completing a request form. If the Data Access Committee approves the request, a material and/or data transfer agreement is signed, and the data and biological materials can be provided. ***Abbreviations:*** CSF, cerebrospinal fluid; EHR, electronic health record; PBMCs, peripheral blood mononuclear cells

### 2.6. Data availability

The collected phenotypes and biomaterials are available to both academic and industry researchers upon request. The process for requesting data and biomaterials is as follows: 1) Requestors send an email with a description of the planned scientific project, the type and quantity of the requested data, and their data protection policies to the MMHB to inquire about data availability, and in the same email, they may also propose collaborations with local researchers; 2) the Data Access Committee evaluates the feasibility of the project and provides a cost estimate for sample extraction, processing, and shipment; 3) at this point, requestors provide documentation confirming ethical approval of the proposed project; 4) the MMHB provides a quote, data- and material transfer agreements (DTA and MTA, respectively); and 5) once the DTA and/or MTA is signed by both parties, the biological materials and double-pseudonymized phenotypic and omics data are sent to the requestor. Depending on the legal process, approximately four to six months should be allowed from submission of the project proposal to delivery of the data and/or biomaterials.

## 3. Sample characteristics

Detailed phenotype definitions of the MMHB Sample and Clinical Sample (i.e., the target population of all patients treated at the DPP and IPPG during the same period who were eligible for inclusion in the MMHB but were not included) are provided in the description of the individual analyses and in *Supplementary Table S1*. Qualitative traits were analyzed with non-parametric pairwise Mann-Whitney U tests, and quantitative traits, with χ2 tests. The significance threshold was set at 0.05. The study protocol was approved by the local ethics committee, and the study was performed in accordance with the Declaration of Helsinki. All participants in the MMHB Sample provided written informed consent. The Clinical Sample served as naturalistic comparator: the data was generated in the clinical routine and irreversibly anonymized for further analysis.

### 3.1. The MMHB Sample

Recruitment for the MMHB started on April 11, 2019. As of October 31, 2021, 578 patients and 104 healthy controls had been included in the MMHB. *Table 2* and *Figure 2* provide an overview of the demographic and clinical characteristics of the sample. Participants were more likely to be recruited in an inpatient setting (67.47%) than in the day hospital (3.46%) or outpatient clinic (29.07%). The analysis included all (independent) clinical (ICD-10) diagnoses the patient received during the observational period (April 11, 2019, to October 31, 2021). The most frequent ICD-10 diagnoses were recurrent depressive disorder (38.78%; ICD-10: F33), alcohol-related disorders (19.88%; ICD-10: F10), schizophrenia (19.69%; ICD-10: F20), depressive episode (15.94%; ICD-10: F32), and personality disorders (13.78%; ICD-10: F60). A full list of the diagnoses and the respective frequencies is provided in *Supplementary Table S3*.

**Table 2.**
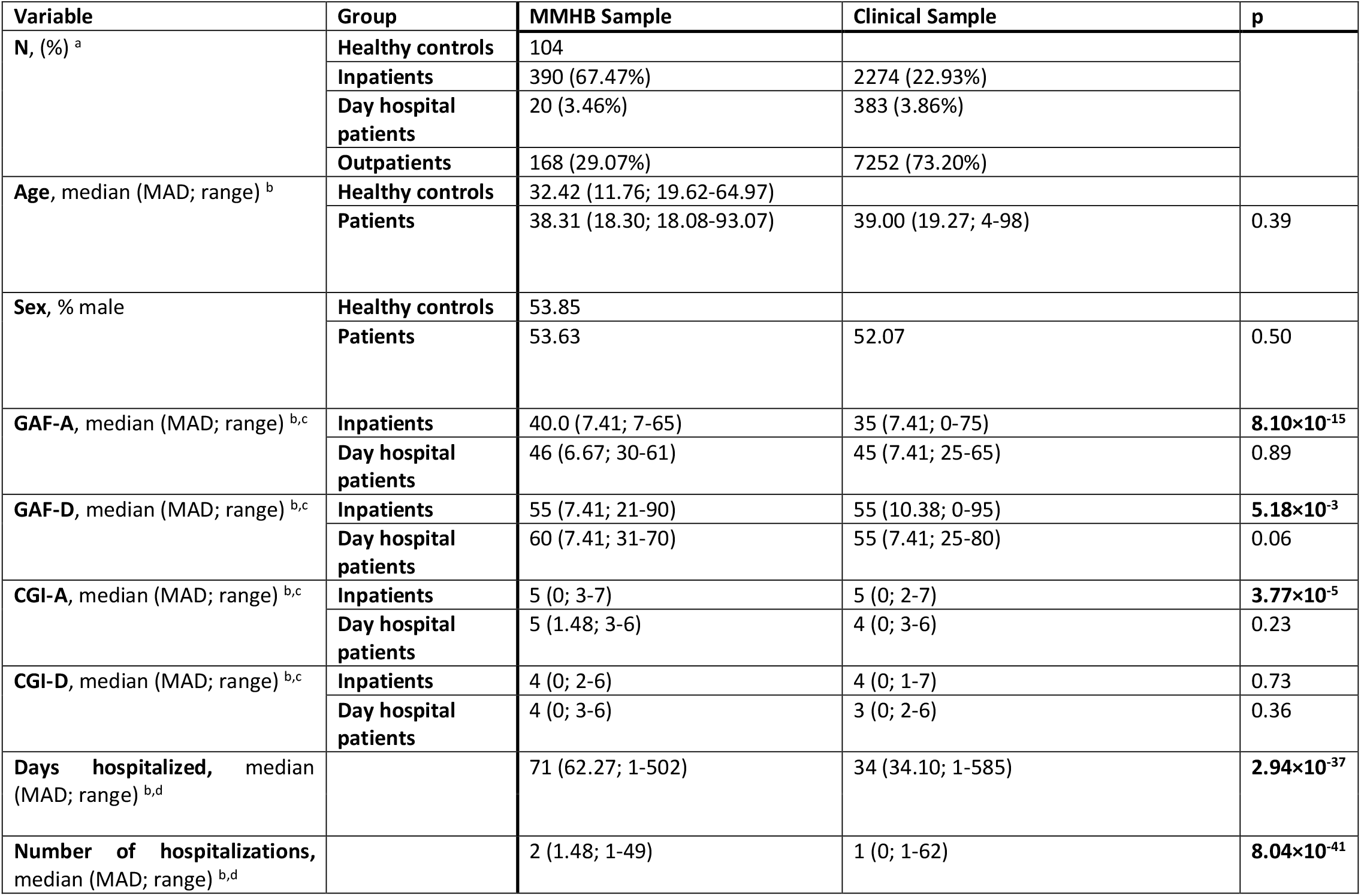
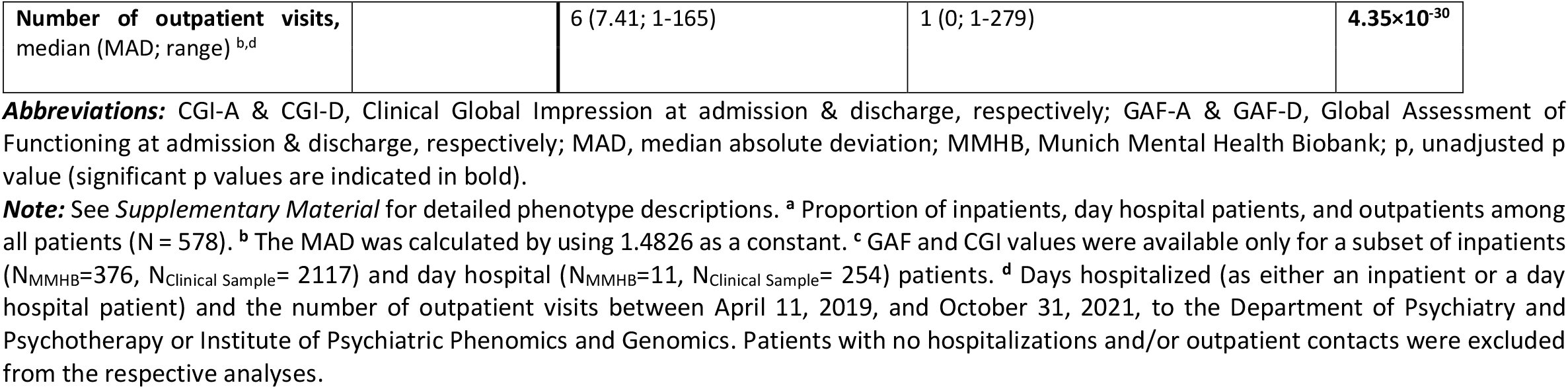
Sample characteristics

**Figure 2:**
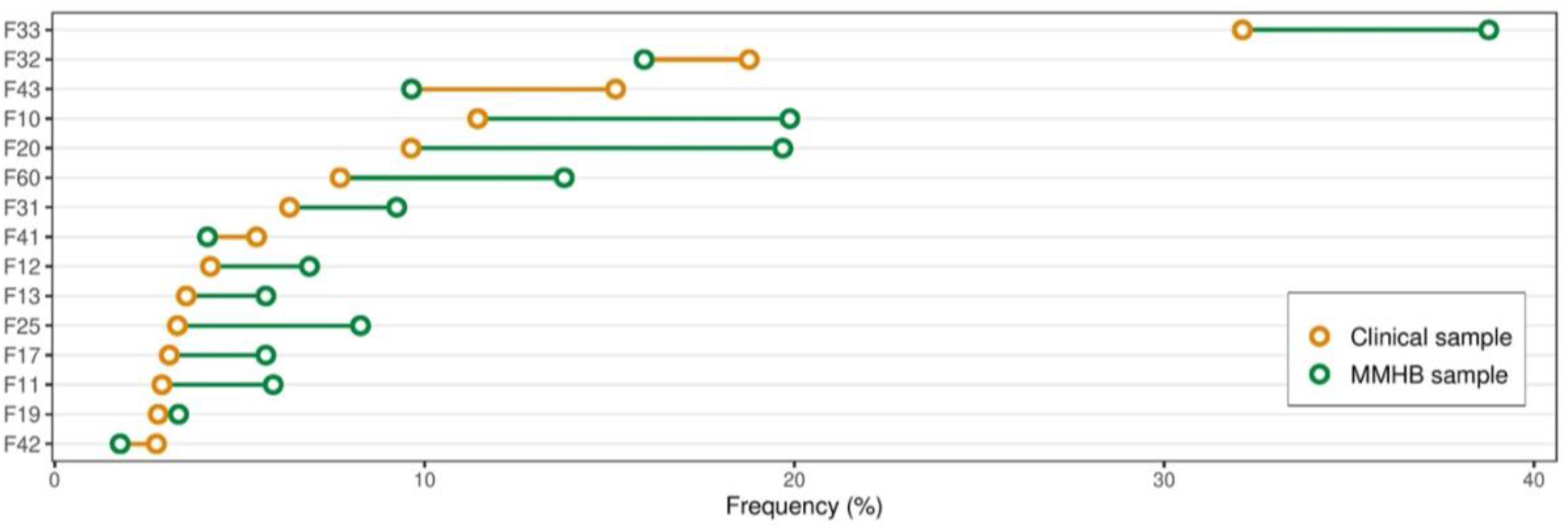
Differences in diagnosis frequency between the Munich Mental Health Biobank (MMHB) and Clinical samples Comparison of the prevalence of the 15 most frequent diagnoses derived from the Clinical Sample in the Munich Mental Health Biobank and the Clinical Sample. The diagnoses were defined as all (independent) clinical (ICD-10) diagnoses the patient received during the observational period (April 11, 2019, to October 31, 2021). A full list of diagnoses and their respective frequencies is provided in *Supplementary Table S3*. ***Abbreviations***: F33, recurrent depressive disorder; F32, depressive episode; F43, adjustment disorder; F10, alcohol-related disorders; F20, schizophrenia; F60, personality disorders; F31, bipolar disorder; F41, other anxiety disorders; F12, mental and behavioural disorders due to use of cannabinoids; F13, mental and behavioural disorders due to use of sedatives or hypnotics; F25, schizoaffective disorder; F17, mental and behavioural disorders due to use of tobacco; F11, mental and behavioural disorders due to use of opioids; F19, mental and behavioural disorders due to multiple drug use and use of other psychoactive substances; F42, obsessive-compulsive disorder; MMHB, Munich Mental Health Biobank

### 3.2. Comparison of the MMHB Sample and the Clinical Sample

Next, we compared the basic sociodemographic and clinical characteristics of the MMHB Sample with the Clinical Sample (i.e., the target population of all patients treated at the DPP and IPPG during the same period who were eligible for inclusion in the MMHB but were not included). The two samples had similar age and sex distributions *(Table 2)*. Clinicians at the DPP and IPPG routinely assess the seven-day functioning and disease severity of inpatients and day hospital patients at admission and discharge by using the Global Assessment of Functioning (GAF) and Clinical Global Impression (CGI) scales, respectively (21,22). According to these measures, patients in both the MMHB and Clinical samples had severe symptoms and major impairments in functioning at admission and moderate symptoms and improved functioning at discharge. However, compared with the Clinical Sample, MMHB participants who had been recruited as inpatients had significantly higher functioning at both admission (GAF-A_MMHB_= 40 vs. GAF-A_Clinical Sample_= 35, Mann-Whitney-U-Test, *p* = 8.10×10^−15^) and discharge (GAF-D_MMHB_= 55 vs. GAF-D_Full Clinical Sample_ = 55, *p* = 5.18×10^−3^) and less severe symptoms at admission (CGI-A_MMHB_= 5 vs. CGI-A_Clinical Sample_= 5, *p* = 3.77×10^−5^). Symptom severity at discharge (CGI-D) was similar in both groups. No significant differences were observed between the MMHB and Clinical samples of day hospital patients. GAF and CGI values were not available for the outpatients. During the observation period, patients in the MMHB Sample were hospitalized more frequently than those in the Clinical Sample (number of hospitalizations: 2 vs. 1; Mann-Whitney U test, *p* = 8.04×10^−41^), for longer periods of time (total length of stay, 71 vs. 37 days; *p* = 2.94×10^−37^), and had more outpatient contacts (6 vs. 1; *p* = 4.35×10^−30^) *(Table 2)*.

*Figure 2* provides an overview of the prevalence in the two samples of the 15 most frequent diagnoses derived from the Clinical Sample. In the MMHB Sample vs the Clinical Sample, we observed the most pronounced differences in the prevalence of recurrent depressive disorder (38.78% vs 32.12%; ICD-10: F33), adjustment disorder (9.65% vs 15.17%; ICD-10: F43), alcohol-related disorders (19.88% vs 11.44%; ICD-10: F10), schizophrenia (19.69% vs 9.64%; ICD-10: F20), personality disorders (13.78% vs 7.72%; ICD-10: F60), and schizoaffective disorder (8.27% vs 3.32%; ICD-10: F25). A full list of diagnoses and the respective frequencies is provided in *Supplementary Table S3*.

## 4. Discussion

The MMHB is a mental health-specific biobank that aims to integrate multimodal research and real-world phenotypic information with biosamples of patients with mental health disorders and study participants, including healthy controls, to facilitate translatable mental health research and, ultimately, improve patient care. Here, we outlined the information technology and laboratory infrastructure and the administrative processes of the MMHB, described the clinical and sociodemographic characteristics of the 578 patients and 104 healthy controls included in the MMHB so far and compared this sample with the overall target population treated at the recruitment facilities. This initial evaluation allows us to discuss the representativeness and scope of the MMHB sample, identify limitations of our current recruitment strategy, and derive action points that could further improve the quality of the MMHB data.

### 4.1. Representativeness

There is increasing evidence that individuals with certain non-random characteristics, such as lower socioeconomic and education status, poorer health, non-European ancestry, and increased cumulative genetic burden for schizophrenia, neuroticism, and attention-deficit hyperactivity disorder, are less likely to participate in clinical studies and more likely to drop out during the follow-up period (9,23–25). These data are concerning because non-participation and attrition not only are associated with a loss of statistical power, but, if non-random, can also influence sample representativeness and thereby bias the generalizability and real-life utility of research findings, health policy decision-making, and ultimately, the equity of health care provision (26). Compared with patients treated at the DPP and IPPG since April 11, 2019 who were not included in the MMHB (Clinical Sample), we found that certain mental health diagnoses, such as schizophrenia, schizoaffective disorder, alcohol-related disorders, and personality disorders, are overrepresented in the MMHB sample, whereas adjustment disorders, for example, are relatively underrepresented, most likely because recruitment so far has been propagated by study teams interested in answering specific research questions. Interestingly, we found evidence that the frequency and length of contact with the health care system differed between the target population (i.e., those not included in the biobank) and the MMHB sample: MMHB participants were not only more frequently hospitalized (median number of hospitalizations in MMHB Sample vs Clinical Sample, 2 vs 1), but they also spent more days in hospital (median, 71 vs 37 days) and had more outpatient appointments (6 vs 1) during the 30-month observational period. The proportion of participants recruited in an inpatient setting was also higher in the MMHB Sample than in the Clinical Sample (67.47% vs 22.93%). These differences could be explained by potential systematic sampling biases in that study personnel and clinicians are more likely to ask “known” patients to consent to being included in the MMHB and/or that patients with repeated contacts with the same institution are more likely to trust that institution and participate in its research efforts. Also, longer hospitalizations and more frequent visits in general represent more opportunities to recruit participants. These observations emphasize the importance of interpersonal factors (such as trust, rapport, and compliance) and the type and frequency of contact with the health care system in study participation. These factors are potential sources of recruitment bias that should be taken into consideration when analyzing the MMHB Sample. Interestingly, despite the more frequent health care utilization, the MMHB Sample had less severe symptoms and higher levels of functioning (measured with CGI and GAF, respectively) at inclusion in the biobank than the Clinical Sample in general. Although this difference is likely not clinically meaningful (the difference in functioning was within a 10-point GAF range), it still suggests that patients who were successfully recruited had better overall social, occupational, and psychological functioning.

### 4.2. Availability of real-world clinical data

As outlined, the current MMHB phenotyping battery includes detailed sociodemographic and self-reported transdiagnostic information that enables research across multiple disorders and patient subgroups. However, like most other mental health-specific biobanks, currently the MMHB includes only a limited set of features from routine clinical care (patient age, sex, zip code, GAF, CGI, diagnosis, and laboratory results). This limited information is a direct result of the limitations of current electronic health record systems, which are markedly restricted in their ability to present relevant mental health information in a standardized, structured, interoperable, and thus machine-readable format. Therefore, our current data mainly include only cross-sectional information and lack real-world clinical data and longitudinal information on disease trajectories. These data modalities, however, represent a main target of current data-driven precision medicine initiatives, which aim at developing reliable, generalizable prediction models of important mental health-related outcomes, such as antidepressant response, suicide attempts, and the transition from clinical high-risk states to psychosis (27–29).

### 4.3. Outlook

A key strength of the MMHB is its close integration into the clinical and scientific infrastructure of a large mental health care center, which not only ensures the necessary domain knowledge for collecting high-quality mental health-specific phenotypes, but also provides direct access to the more than 3000 individual patients treated annually at the DPP and IPPG and thus a strong growth potential (*Figure 3*). Moreover, this target population has good sociodemographic representativeness because Germany has universal health insurance, ensuring that treatment is offered to all individuals in need. A further asset of the MMHB is the wide age range of eligible participants and the diversity of diagnoses, which enables the study of all stages of the disease trajectory across the whole lifespan and diagnostic spectrum.

**Figure 3:**
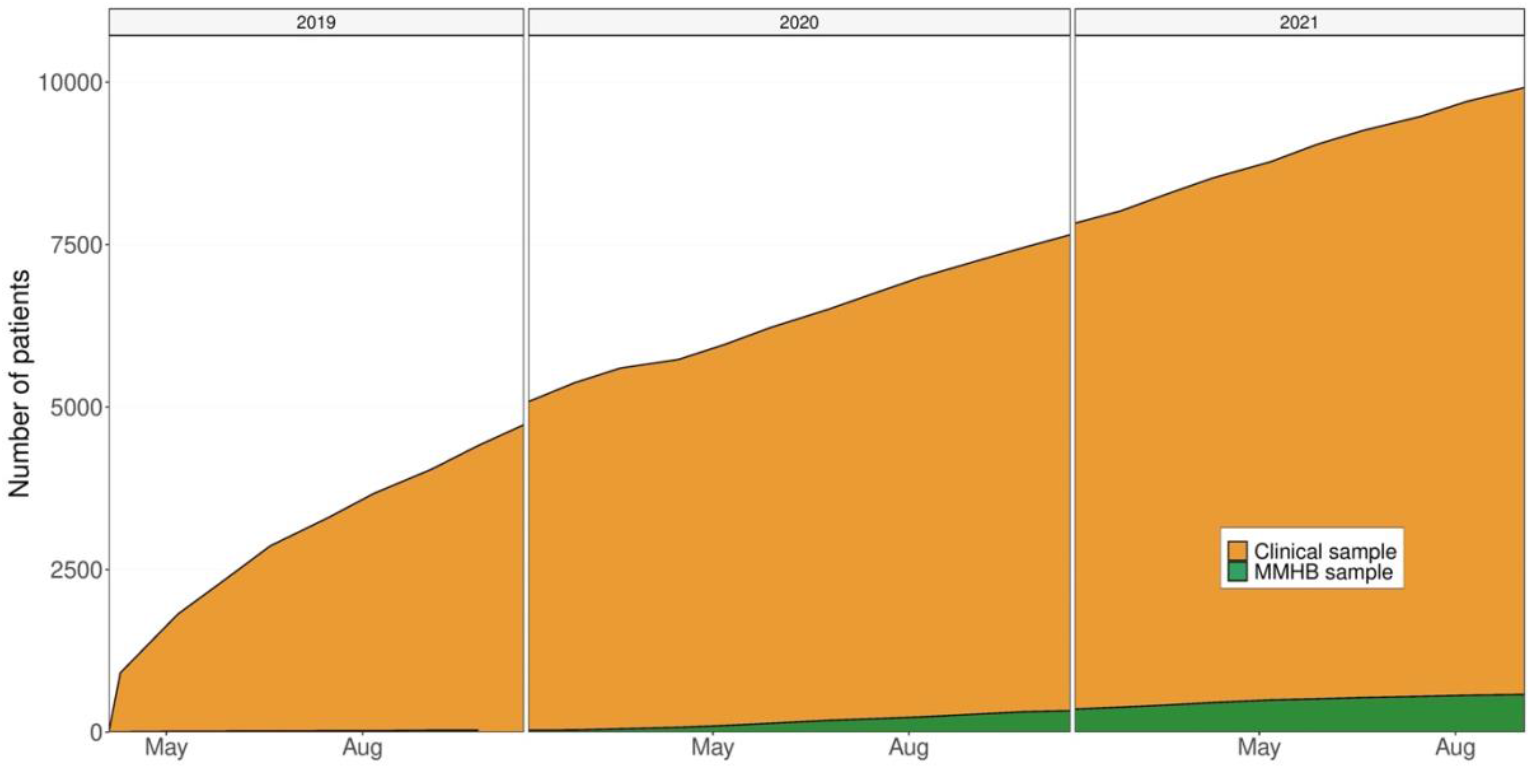
Growth of the Munich Mental Health Biobank (MMHB) and Clinical samples over time ***Abbreviation:*** MMHB, Munich Mental Health Biobank

Limitations of the MMHB include the observed differences between the MMHB and Clinical samples, the current sparsity of available real-world data, and the moderate recruitment pace (only 5.56% of the 10,494 patients who were treated at the DPP and IPPG during the 30-month observational period and were thus eligible for inclusion were recruited into the MMHB, *Figure 3*).

To address these limitations, in the next phase of our biobanking effort, we will move from active, interest-driven biobank recruitment towards a more passive, automatized participatory system. To achieve this, we will first introduce the Broad Consent of the German Medical Informatics Initiative, for which we have developed an additional Psychiatry Module tailored to the needs of mental health research (30). This modified Broad Consent, which will be valid for 5 years, will enable the scientific use of prospectively collected routine clinical and insurance data, and, via the Psychiatry Module, additionally collected mental health-specific phenotypes, magnetic resonance imaging, electroencephalogram data, and biosamples. Next, we will introduce the Clinical Phenotyping Platform (CliPP), a software solution that enables the collection of structured and standardized (Fast Healthcare Interoperability Resource (FHIR) format) routine mental health data directly from patients and health care providers. Owing to the Broad Consent and the CliPP, the consent process and the phenotypic assessments will be incorporated into routine admission procedures. Their introduction will broaden the available data modalities, reduce the time spent on patient recruitment, and decrease the risk of sampling bias due to physician and/or patient preferences. Furthermore, we will continuously monitor the representativeness of the MMHB Sample, analyze the factors that influence biobank participation, and, if necessary, revise study design and recruitment strategies. Through these measures, we will facilitate recruitment of participants into the MMHB, improve clinician and patient commitment and the representativeness of our sample, and ultimately, increase the scientific and clinical value of the MMHB Sample. Large future transdiagnostic cohorts from the MMHB will leverage forward and reverse translational research in an informative framework spanning preclinical models via disorder focused clinical trials to real-world applications in a transdiagnostic spectrum of mental health disorders including their developmental dynamics and co-morbidities.

## Data Availability

All data stored in the Munich Mental Health Biobank are available upon reasonable request to the authors.

## References

1. Liu Q, He H, Yang J, Feng X, Zhao F, Lyu J. Changes in the global burden of depression from 1990 to 2017: Findings from the Global Burden of Disease study. J Psychiatr Res. 2020 Jul 1;126:134–40.

2. John A, McGregor J, Fone D, Dunstan F, Cornish R, Lyons RA, et al. Case-finding for common mental disorders of anxiety and depression in primary care: an external validation of routinely collected data. BMC Med Inform Decis Mak. 2016 Mar 15;16:35.

3. Biobanking.com. Large Study on Mental Health Genetics Launched by Estonian Biobank [Internet]. Biobanking.com. 2021 [cited 2022 Apr 12]. Available from: https://www.biobanking.com/large-study-on-mental-health-genetics-launched-by-estonian-biobank/

4. Davis K, Hotopf M. Mental health phenotyping in UK Biobank. Prog Neurol Psychiatry. 2019;23(1):4–7.

5. Gehring S, Eulenfeld R. German Medical Informatics Initiative: Unlocking Data for Research and Health Care. Methods Inf Med. 2018 Jul;57(Suppl 1):e46–9.

6. Witt S, Dukal H, Hohmeyer C, Radosavljevic-Bjelic S, Schendel D, Frank J, et al. Biobank of Psychiatric Diseases Mannheim – BioPsy. Open J Bioresour. 2016 Jul 8;3(1):e2.

7. Klingler C, von Jagwitz-Biegnitz M, Hartung ML, Hummel M, Specht C. Evaluating the German Biobank Node as Coordinating Institution of the German Biobank Alliance: Engaging with Stakeholders via Survey Research. Biopreservation Biobanking. 2020 Apr;18(2):64–72.

8. Hummel M, Specht C. Biobanks for future medicine. J Lab Med. 2019 Dec 1;43(6):383–8.

9. Budde M, Anderson-Schmidt H, Gade K, Reich-Erkelenz D, Adorjan K, Kalman JL, et al. A longitudinal approach to biological psychiatric research: The PsyCourse study. Am J Med Genet Part B Neuropsychiatr Genet Off Publ Int Soc Psychiatr Genet. 2019 Mar;180(2):89–102.

10. Bush K, Kivlahan DR, McDonell MB, Fihn SD, Bradley KA. The AUDIT alcohol consumption questions (AUDIT-C): an effective brief screening test for problem drinking. Ambulatory Care Quality Improvement Project (ACQUIP). Alcohol Use Disorders Identification Test. Arch Intern Med. 1998 Sep 14;158(16):1789–95.

11. Grabe HJ, Schulz A, Schmidt CO, Appel K, Driessen M, Wingenfeld K, et al. [A brief instrument for the assessment of childhood abuse and neglect: the childhood trauma screener (CTS)]. Psychiatr Prax. 2012 Apr;39(3):109–15.

12. Smith BW, Dalen J, Wiggins K, Tooley E, Christopher P, Bernard J. The brief resilience scale: assessing the ability to bounce back. Int J Behav Med. 2008;15(3):194–200.

13. Hughes ME, Waite LJ, Hawkley LC, Cacioppo JT. A Short Scale for Measuring Loneliness in Large Surveys: Results From Two Population-Based Studies. Res Aging. 2004;26(6):655–72.

14. Lubben J, Blozik E, Gillmann G, Iliffe S, von Renteln Kruse W, Beck JC, et al. Performance of an abbreviated version of the Lubben Social Network Scale among three European community-dwelling older adult populations. The Gerontologist. 2006 Aug;46(4):503–13.

15. Topp CW, Østergaard SD, Søndergaard S, Bech P. The WHO-5 Well-Being Index: a systematic review of the literature. Psychother Psychosom. 2015;84(3):167–76.

16. Kroenke K, Spitzer RL, Williams JB. The PHQ-9: validity of a brief depression severity measure. J Gen Intern Med. 2001 Sep;16(9):606–13.

17. Roenneberg T, Wirz-Justice A, Merrow M. Life between clocks: daily temporal patterns of human chronotypes. J Biol Rhythms. 2003 Feb;18(1):80–90.

18. Barton BB, Zagler A, Engl K, Rihs L, Musil R. Prevalence of obesity, metabolic syndrome, diabetes and risk of cardiovascular disease in a psychiatric inpatient sample: results of the Metabolism in Psychiatry (MiP) Study. Eur Arch Psychiatry Clin Neurosci. 2020 Aug;270(5):597–609.

19. Cappelleri JC, Bushmakin AG, Gerber RA, Leidy NK, Sexton CC, Lowe MR, et al. Psychometric analysis of the Three-Factor Eating Questionnaire-R21: results from a large diverse sample of obese and non-obese participants. Int J Obes 2005. 2009 Jun;33(6):611–20.

20. Rek SV, Bühner M, Reinhard MA, Freeman D, Keeser D, Adorjan K, et al. The COVID-19 Pandemic Mental Health Questionnaire (CoPaQ): psychometric evaluation and compliance with countermeasures in psychiatric inpatients and non-clinical individuals. BMC Psychiatry. 2021 Aug 31;21(1):426.

21. Guy W. ECDEU assessment manual for psychopharmacology. US Department of Health, Education, and Welfare, Public Health Service …; 1976.

22. Jones SH, Thornicroft G, Coffey M, Dunn G. A brief mental health outcome scalereliability and validity of the Global Assessment of Functioning (GAF). Br J Psychiatry J Ment Sci. 1995 May;166(5):654–9.

23. Pirastu N, Cordioli M, Nandakumar P, Mignogna G, Abdellaoui A, Hollis B, et al. Genetic analyses identify widespread sex-differential participation bias. Nat Genet. 2021 May 1;53(5):663–71.

24. Taylor AE, Jones HJ, Sallis H, Euesden J, Stergiakouli E, Davies NM, et al. Exploring the association of genetic factors with participation in the Avon Longitudinal Study of Parents and Children. Int J Epidemiol. 2018 Aug 1;47(4):1207–16.

25. Tyrrell J, Zheng J, Beaumont R, Hinton K, Richardson TG, Wood AR, et al. Genetic predictors of participation in optional components of UK Biobank. Nat Commun. 2021 Feb 9;12(1):886.

26. Larsson H. The importance of selection bias in prospective birth cohort studies. JCPP Adv. 2021;1(3):e12043.

27. Koutsouleris N, Dwyer DB, Degenhardt F, Maj C, Urquijo-Castro MF, Sanfelici R, et al. Multimodal Machine Learning Workflows for Prediction of Psychosis in Patients With Clinical High-Risk Syndromes and Recent-Onset Depression. JAMA Psychiatry. 2021 Feb 1;78(2):195–209.

28. Barak-Corren Y, Castro VM, Nock MK, Mandl KD, Madsen EM, Seiger A, et al. Validation of an Electronic Health Record-Based Suicide Risk Prediction Modeling Approach Across Multiple Health Care Systems. JAMA Netw Open. 2020 Mar 2;3(3):e201262.

29. Chekroud AM, Zotti RJ, Shehzad Z, Gueorguieva R, Johnson MK, Trivedi MH, et al. Crosstrial prediction of treatment outcome in depression: a machine learning approach. Lancet Psychiatry. 2016 Mar;3(3):243–50.

30. Semler SC, Wissing F, Heyder R. German Medical Informatics Initiative. Methods Inf Med. 2018 Jul;57(Suppl 1):e50–6.

