## Supplementary Material for "A blueprint for biobanking in everyday clinical practice in psychiatry: The Munich Mental Health Biobank"

### Supplementary Table S1: Phenotype definitions

|  | **Munich Mental Health Biobank Sample** | **Clinical Sample** |
| --- | --- | --- |
| **Age** | Age at inclusion in biobank | Age at first clinical presentation (either as out- or inpatient or day hospital patient) after April 11, 2019 |
| **GAF-A** | Global Assessment of Functioning (1) at admission: Seven-day functioning assessed by a clinician at admission | |
| **GAF-D** | Global Assessment of Functioning at discharge: Seven-day functioning assessed by a clinician at discharge | |
| **CGI-A** | Clinical Global Impression (2) at admission: Seven-day disease severity assessed by a clinician at admission | |
| **CGI-D** | Clinical Global Impression at discharge: Seven-day disease severity assessed by a clinician at discharge | |
| **Diagnosis** | Clinical (ICD-10) diagnoses the patient received between April 11, 2019, and October 31, 2021, in the DPP and IPPG | |
| **Days hospitalized** | Number of days spent in the DPP as an inpatient or day hospital patient between April 11, 2019, and October 31, 2021 | |
| **Number of hospitalizations** | Number of hospitalizations in the DPP as an inpatient or day hospital patient between April 11, 2019, and October 31, 2021 | |
| **Number of outpatient contacts** | Number of outpatient contacts (on different days) in the DPP and/or IPPG between April 11, 2019, and October 31, 2021 | |

DPP, Department of Psychiatry and Psychotherapy; IPPG, Institute of Psychiatric Phenomics and Genomics. Both the DPP and IPPG are part of the hospital system of the University of Munich (LMU) in Munich, Germany.

### Supplementary Table S2: Overview of the collected biosamples

| **Biosample** | **Supplier** | **CatNo** | **Purpose** | **Storage** |
| --- | --- | --- | --- | --- |
| Pax Gene blood RNA tube | BD | 762165 | RNA | -80°C |
| 7,5 ml K3EDTA Monovette | Sarstedt | 01.1605.001 | DNA | -80°C |
| 9 ml K3EDTA Monovette | Sarstedt | 02.1066.001 | Plasma | -80°C |
| 9 ml Serum S-Monovette | Sarstedt | 02.1063.001 | Serum | -80°C |
| BD Vacutainer 10ml Glass Sodium Heparin Tubes | BD | 368480 | PBMC | -176°C |

### Supplementary Table S3: Full list of all (independent) clinical (ICD-10) diagnoses made in patients in the Munich Mental Health Biobank (MMHB) and Clinical samples during the observational period (April 11, 2019, to October 31, 2021)

| **Diagnosis** | **ICD-10** | **Prevalence,** % | | **Difference,** % |
| --- | --- | --- | --- | --- |
|  |  | **Munich Mental Health Biobank Sample** | **Clinical Sample** |  |
| Recurrent depressive disorder | F33 | 38.78 | 32.12 | 6.66 |
| Depressive Episode | F32 | 15.94 | 18.78 | 2.84 |
| Adjustment disorder | F43 | 9.65 | 15.17 | 5.52 |
| Mental and behavioural disorders due to use of alcohol | F10 | 19.88 | 11.44 | 8.44 |
| Schizophrenia | F20 | 19.69 | 9.64 | 10.05 |
| Personality disorders | F60 | 13.78 | 7.72 | 6.06 |
| Bipolar disorder | F31 | 9.25 | 6.35 | 2.9 |
| Other anxiety disorders | F41 | 4.13 | 5.46 | 1.33 |
| Mental and behavioural disorders due to use of cannabinoids | F12 | 6.89 | 4.21 | 2.68 |
| Mental and behavioural disorders due to use of sedatives or hypnotics | F13 | 5.71 | 3.56 | 2.15 |
| Schizoaffective disorder | F25 | 8.27 | 3.32 | 4.95 |
| Mental and behavioural disorders due to use of tobacco | F17 | 5.71 | 3.1 | 2.61 |
| Mental and behavioural disorders due to use of opioids | F11 | 5.91 | 2.9 | 3.01 |
| Mental and behavioural disorders due to multiple drug use and use of other psychoactive substances | F19 | 3.35 | 2.8 | 0.55 |
| Obsessive-compulsive disorder | F42 | 1.77 | 2.75 | 0.98 |
| Other mental disorders due to brain damage and dysfunction and to physical disease | F06 | 1.77 | 2.58 | 0.81 |
| Tic disorders | F95 | 0.39 | 2.36 | 1.97 |
| Somatoform disorders | F45 | 0.98 | 2.29 | 1.31 |
| Acute and transient psychotic disorders | F23 | 1.97 | 2.08 | 0.11 |
| Persistent mood [affective] disorders | F34 | 2.17 | 1.91 | 0.26 |
| Phobic anxiety disorders | F40 | 2.76 | 1.74 | 1.02 |
| Hyperkinetic disorders | F90 | 0.79 | 1.61 | 0.82 |
| Pervasive developmental disorders | F84 | 1.18 | 1.29 | 0.11 |
| Eating disorders | F50 | 1.57 | 1.26 | 0.31 |
| Mental and behavioural disorders due to use of cocaine | F14 | 2.95 | 1.24 | 1.71 |
| Mental and behavioural disorders due to use of other stimulants, including caffeine | F15 | 1.18 | 1.17 | 0.01 |
| Mixed and other personality disorders | F61 | 1.57 | 1.09 | 0.48 |
| Persistent delusional disorders | F22 | 0.2 | 0.84 | 0.64 |
| Dissociative [conversion] disorders | F44 | 1.57 | 0.74 | 0.83 |
| Habit and impulse disorders | F63 | 0.39 | 0.54 | 0.15 |
| Mild mental retardation | F70 | 0.2 | 0.4 | 0.2 |
| Unspecified dementia | F03 | 0.39 | 0.39 | 0 |
| Delirium, not induced by alcohol and other psychoactive substances | F05 |  | 0.28 |  |
| Gender identity disorders | F64 | 0.2 | 0.27 | 0.07 |
| Mental and behavioural disorders due to use of hallucinogens | F16 | 0.2 | 0.25 | 0.05 |
| Nonorganic sleep disorders | F51 | 0.2 | 0.25 | 0.05 |
| Other neurotic disorders | F48 |  | 0.21 |  |
| Schizotypal disorder | F21 |  | 0.19 |  |
| Mental and behavioural disorders associated with the puerperium, not elsewhere classified | F53 |  | 0.19 |  |
| Manic episode | F30 |  | 0.18 |  |
| Vascular dementia | F01 |  | 0.15 |  |
| Personality and behavioural disorders due to brain disease, damage and dysfunction | F07 | 0.2 | 0.13 | 0.07 |
| Specific developmental disorders of speech and language | F80 | 0.2 | 0.09 | 0.11 |
| Unspecified nonorganic psychosis | F29 | 0.2 | 0.08 | 0.12 |
| Sexual dysfunction, not caused by organic disorder or disease | F52 |  | 0.07 |  |
| Unspecified disorder of adult personality and behaviour | F69 |  | 0.07 |  |
| Conduct disorder | F91 |  | 0.07 |  |
| Emotional disorders with onset specific to childhood | F93 |  | 0.07 |  |
| Mental and behavioural disorders due to use of volatile solvents | F18 | 0.39 | 0.06 | 0.33 |
| Abuse of non-dependence-producing substances | F55 |  | 0.06 |  |
| Other disorders of adult personality and behaviour | F68 |  | 0.06 |  |
| Moderate mental retardation | F71 |  | 0.06 |  |
| Other behavioural and emotional disorders with onset usually occurring in childhood and adolescence | F98 | 0.39 | 0.06 | 0.33 |
| Induced delusional disorder | F24 |  | 0.05 |  |
| Mixed disorders of conduct and emotions | F92 |  | 0.05 |  |
| Enduring personality changes, not attributable to brain damage and disease | F62 | 0.2 | 0.04 | 0.16 |
| Unspecified mental retardation | F79 |  | 0.04 |  |
| Specific developmental disorders of scholastic skills | F81 |  | 0.04 |  |
| Specific developmental disorder of motor function | F82 |  | 0.04 |  |
| Mental disorder, not otherwise specified | F99 |  | 0.04 |  |
| Organic amnesic syndrome, not induced by alcohol and other psychoactive substances | F04 |  | 0.02 |  |
| Other nonorganic psychotic disorders | F28 |  | 0.02 |  |
| Other mood [affective] disorders | F38 |  | 0.02 |  |
| Psychological and behavioural factors associated with disorders or diseases classified elsewhere | F54 |  | 0.02 |  |
| Mixed specific developmental disorders | F83 |  | 0.02 |  |
| Disorders of social functioning with onset specific to childhood and adolescence | F94 |  | 0.02 |  |
| Dementia in Alzheimer disease | F00 |  | 0.01 |  |
| Dementia in other diseases classified elsewhere | F02 |  | 0.01 |  |
| Unspecified organic or symptomatic mental disorder | F09 |  | 0.01 |  |
| Disorders of sexual preference | F65 |  | 0.01 |  |
| Psychological and behavioural disorders associated with sexual development and orientation | F66 |  | 0.01 |  |
| Dissociated Intelligence | F74 |  | 0.01 |  |

### Supplementary References

1.	Jones SH, Thornicroft G, Coffey M, Dunn G. A brief mental health outcome scale-reliability and validity of the Global Assessment of Functioning (GAF). Br J Psychiatry. 1995 May;166(5):654–9.

2.	Guy W. ECDEU assessment manual for psychopharmacology. US Department of Health, Education, and Welfare, Public Health Service …; 1976.
